# Impact of an over-the-counter “sleep lotion” on salivary melatonin levels and sleep quality: a randomized controlled trial

**DOI:** 10.1101/2023.10.16.23297058

**Authors:** Clairissa Ponce, Amanda D. Razon, Joey Chao, Sydney K. Nakagawa, Megan M. Peterson, Angelina Y. Roque, Maya A. Vanderpool, Michael J. Ferracane, Lisa E. Olson

## Abstract

Many over-the-counter products such as bubble baths, room sprays, and lotions claim they contain the pineal hormone melatonin and promote sleep. In this randomized, controlled, double-blind crossover trial we compared the impact of a commercial “sleep lotion” versus a placebo control lotion. Our sample of 63 undergraduate college students scored an average of 6.3 ± 2.7 on the Global Pittsburgh Sleep Quality Index, with 65% above the cutoff of 5 indicating poor sleep quality in the previous 30 days. Participants applied lotions on two different nights, one hour before bedtime. To assess whether melatonin was absorbed through the skin and circulating systemically, we conducted enzyme linked immunosorbent assays on saliva samples to quantitate melatonin levels. We also assessed sleep quality the night after lotion application with a modified Pittsburgh Sleep Quality Index. The melatonin-containing sleep lotion dramatically impacted salivary melatonin levels, increasing them up to ∼1000 fold compared to the placebo control lotion. Sleep quality in the overall group was not impacted by the lotion, but in a subsample of the poor sleepers, the lotion improved sleep quality. High Performance Liquid Chromatography of the sleep lotion revealed the presence of 2.4 ± 0.1 mg melatonin/g lotion, or a 0.24 ± 0.01% formulation. Caution should be taken by consumers using over-the-counter melatonin lotions because the undisclosed dosage is high and well absorbed by the skin. Clinicaltrials.gov ID NCT06053385

## Introduction

Regulated amounts of melatonin have been shown to improve sleep quality in adults (Fatemeh et al., 2022; Xie et al., 2017) and adolescents (Eckerberg, Lowden, Nagai, & Akerstedt, 2012) with certain sleep disorders, and the use of melatonin in the United States has dramatically increased in the last decade (Li, Somers, Xu, Lopez-Jimenez, & Covassin, 2022). Unlike the United Kingdom, Europe, Japan, Australia and Canada where it must be prescribed by physicians (Grigg-Damberger & Ianakieva, 2017), melatonin is available over-the-counter in the United States, and it is not classified as a drug by the Federal Drug Administration (Skrzelowski, Brookhaus, Shea, & Berlau, 2021). In addition to oral supplements, melatonin can be purchased in the form of room sprays, lotions, bubble baths, and patches for which the efficacy is unknown. “Dr. Teal’s” (PDC Brands, Stamford, CT) is a popular maker of melatonin-containing aromatherapies, lotions, and bath products that claim to “calm the mind for a good night’s sleep” (https://www.drteals.com/products/). In 2018, Dr. Teal’s was the leading brand for bath fragrance/bubble bath in the United States with $104 million USD in sales; this was over three times more than the next closest competitor (Statista, 2022).

College students often have poor sleep quality (Afandi et al., 2013; Hagedorn et al., 2021; Lund, Reider, Whiting, & Prichard, 2010; Peltz, Bodenlos, Kingery, & Rogge, 2021), which can be associated with depression, anxiety, and stress (Becker et al., 2018; Choueiry et al., 2016; Zou et al., 2020). Both too short and too long of sleep duration is predictive of poorer academic performance (Bermudez et al., 2022). In this study, we aim to measure whether melatonin can be absorbed through the skin from Dr. Teal’s Sleep Lotion and if it has an impact on sleep in an undergraduate population.

## Methods

This randomized, controlled, double-blind crossover trial (Figure 1) was approved by our Institutional Review Board (approval number 2021-37-REDLANDS) and registered with clinicaltrials.gov (NCT06053385). Participants provided informed consent, and the experiment was performed according to the Declaration of Helsinki. Our intended sample size of 62 participants was determined with a power calculation (Faul, Erdfelder, Lang, & Buchner, 2007) using the results of pilot data, estimates of variability from previous work (Burgess & Fogg, 2008), and 95% sensitivity to detect a moderate effect size. Participants were undergraduate students at a small liberal arts university in California, USA between the ages of 18-24.

**Figure 1.**
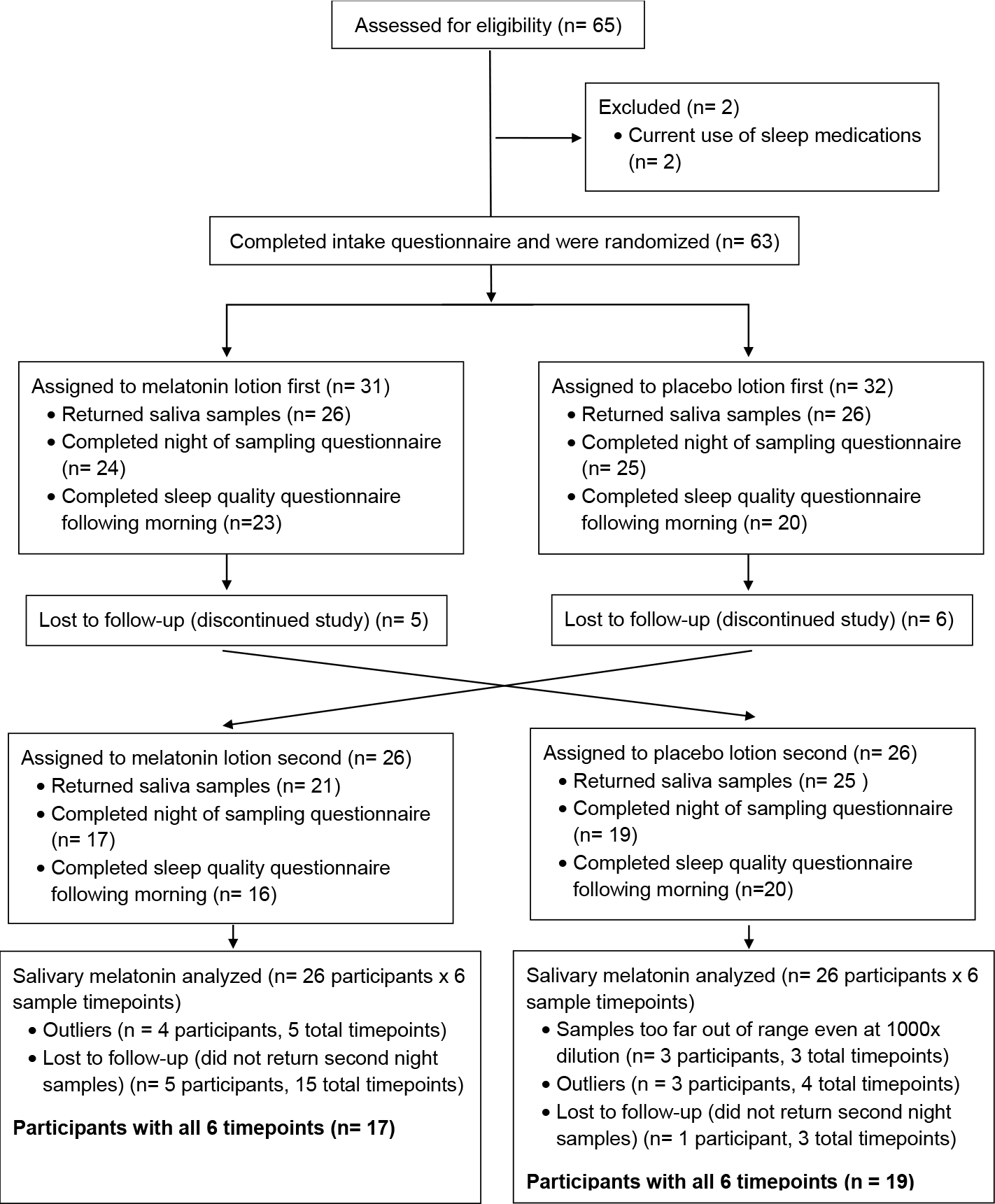
Consolidated Standards of Reporting Trials (CONSORT) diagram for the study.

Exclusion criteria included pregnancy, use of sleep medications in the previous two weeks, and allergies/sensitivity to scented lotion. If participants completed the study, they received a wellness gift (such as hand sanitizer, body wash, air freshener, or lip balm) worth less than $5. Data collection occurred from February 2022 to February 2023.

After providing consent, participants answered an electronic questionnaire on demographic information; use of caffeine, alcohol, medications, and glasses/contacts; exercise; and the Pittsburgh Sleep Quality Index (PSQI) to measure sleep quality in the last month (Buysse, Reynolds, Monk, Berman, & Kupfer, 1989). The PSQI is scored between 0 – 21, with higher scores indicating worse sleep. Scores above the cutoff of 5 indicate poor sleep.

We tested two lotions that are widely available over-the-counter in pharmacies and discount department stores: Dr. Teal’s Sleep Lotion with Melatonin and Essential Oils and Dr. Teal’s Soothing Lavender Essential Oil Body Lotion as a scent-matched placebo control (PDC Brands, Stamford, CT). Lotion application and saliva sample collection occurred at the participant’s home. Participants applied 7 g of lotion to their hands and arms one hour before bedtime on two separate nights. The order of which lotion was applied on the first versus the second night was randomized and allocated by L.E.O. (Urbaniak & Plous, 2013); both participants and researchers doing the analysis were blinded to the lotion type. These two nights were approximately one week apart; participants were asked to choose days with similar schedules when they could avoid pitted fruits, bananas, and chocolate.

An hour prior to saliva collection, participants were asked to avoid foods with high sugar, acidity, or caffeine, and to not brush their teeth or eat a meal. Participants took their own saliva samples three times on each night: 1) immediately prior to lotion application, 2) 30 minutes following application, and 3) 1 hour following application (at planned bedtime). They were provided a kit containing sample tubes and saliva collection aids for passive drool (Salimetrics, State College, PA) and a QR code to a video demonstration. Samples were frozen until analysis.

The following morning, participants completed an electronic PSQI modified to ask questions about the quality of the previous night’s sleep only. Changing question wording from “How often in the past month have you had trouble sleeping due to…” to “Last night, did you have trouble sleeping due to…” yielded possible scores from 0 to 14 rather than the original 0 to 21.

Reagents for enzyme-linked immunosorbent assays were purchased from Salimetrics (State College, PA). In this assay, sample melatonin competes with melatonin conjugated to horseradish peroxidase, which reacts with tetramethylbenzidine. Optical density at 450 nm and initial estimates for melatonin concentration were assessed using an AgileReader (ACTGene, Piscataway, NJ). Standards from 0.78 pg/mL to 50 pg/mL were used to create a four parameter logistic standard curve of B/B_0_ using mycurvefit.com software for final melatonin concentration values. Participant samples were diluted to 10x, 100x, or 1000x if necessary to fall within the standard curve and all samples were tested in at least duplicate. Undiluted salivary melatonin samples whose absorbances fell below the standard curve (n = 4 samples) were assigned a value of 0.78 pg/ml, the lowest value of the standards. Some salivary melatonin sample absorbances were so far above the standard curve even at 1000-fold dilution that the AgileReader software (ACTGene, Piscataway, NJ) did not generate an extrapolation for concentration; these samples (n = 3) were excluded from analysis. Some samples were above the standard curve but were extrapolated by the AgileReader software; rather than use extrapolated values, these samples (n = 3) were assigned a value equivalent to the next highest melatonin concentration at that timepoint in order to be cautious. The average intraassay coefficient of variation was 3% and interassay coefficient of variation for this study was 18%.

We adapted a protocol to prepare lotion extracts (Usher, Simmons, Keating, & Rossi, 2015) for high performance liquid chromatography (HPLC) analysis. A solution of 0.5 g lotion and 5 mL 20% NaCl (EMD MilliporeSigma, Burlington, MA) was vortexed for 30 seconds and subsequently centrifuged for 6 minutes at 4000 rpm. The aqueous layer was filtered using a syringe disc filter containing a 0.22 μm polyvinylidene difluoride membrane. For recovery studies, melatonin (99% pure, ThermoFisher Scientific, Waltham, MA) was added to lotion samples at 50 μg melatonin/mL solvent prior to beginning the extraction protocol. All extractions were performed in triplicate. Melatonin standards (10 – 100 μg/mL) were prepared in ultrapure water.

HPLC analyses were performed using an Agilent 1220 LC System (Agilent, Santa Clara, CA) equipped with a C18 Prodigy ODS(2) reverse-phase analytical column (Phenomenex, 4.6 x 150 mm, 150 Å) at a flow rate of 1 mL/min. The gradient began at 1 min and was 15 – 35% solvent B over 20 min. Solvent A was ultrapure water containing 0.1% trifluoroacetic acid; solvent B was acetonitrile containing 0.1% trifluoroacetic acid. All standards and lotion samples were analyzed in triplicate using an injection volume of 20 μL, and melatonin quantified using absorbance at 215 nm. Ultrapure water (>18 MΩ-cm) was obtained from a Barnstead E-PURE system (ThermoFisher Scientific, Waltham, MA); HPLC-grade acetonitrile was obtained from VWR (Radnor, PA); trifluoroacetic acid was obtained from ThermoFisher Scientific (Waltham, MA).

All statistical analyses were performed using IBM SPSS statistical software (International Business Machines Corporation, Armonk, NY). Extreme outliers as identified by SPSS (± 3x interquartile range) were removed. Data were transformed by log_10_ or square root (if data set included values of zero) to achieve a normal distribution if necessary; when this was unsuccessful, non-parametric analyses were used. Analyses comparing race (White vs. Non-White), ethnicity (Hispanic/Latino vs. Non-Hispanic/Latino), first generation status (yes vs. no), and gender (male vs. female) were performed with independent samples t-tests or Mann-Whitney-U tests as appropriate with an alpha of 0.05. When multiple tests were conducted on the same dependent variable, the Benjamini-Hochberg correction for multiple testing was used to control the false discovery rate to 0.05 (McDonald, 2014) and only those p values significant after correction are reported. Salivary melatonin levels were compared using a two-way repeated measures ANOVA with an alpha level of 0.05. This 2 x 3 ANOVA tested treatment (within subjects factor: placebo control lotion vs. sleep lotion) and time (within subjects factor: 60 min prior to bedtime/prior to lotion application; 30 min prior to bedtime/30 min after lotion application; and at bedtime/60 min after lotion application). Assumptions of ANOVA were checked and appropriate corrections applied when necessary.

## Results

Of the 63 college students who participated in this study, 28 (44 %) were White, 5 (8%) Asian, 4 (6%) Black or African-American, 1 (2%) American Indian or Alaska Native, 4 (6%) multiracial, 16 (25%) other, and 5 (8%) preferred not to disclose; due to small samples sizes, race was only analyzed by comparing White to Non-White participants, excluding those who preferred not to disclose. More participants were Hispanic or Latino (37/59%) than not (26/41%). Approximately half (46%) of the participants were in the first generation in their family to pursue a four-year college degree (30 first generation; 33 non-first generation). The majority (81%) of participants were female (51 female, 10 male, 1 non-binary/third gender, 1 prefer not to disclose). Due to small sample sizes, gender was analyzed by comparing to males versus females only.

### Baseline variables

The mean sleep quality in the last month as measured by the PSQI was 6.3 ± 2.7; 65% of the participants scored above the Global PSQI cutoff of 5, indicating poor quality sleep. Global PSQI was not different by race, ethnicity, first generation student status, or gender (p’s > 0.05). Global PSQI was better in participants who vigorously exercised more than once per week for at least 30 minutes (5.4 ± 2.3, n = 37) compared to those who did not (7.7 ± 2.7, n = 26, p < 0.001, Cohen’s d = 0.97).

Participants reported that they typically slept 7.1 ± 0.13 hours per night; this was not different by race, ethnicity, first generation student status, or gender (p’s > 0.05). On average, participants went to bed at 00:03 ± 01:11, and this was different by race with White participants going to bed 42 minutes (95% CI 7 – 78 minutes) earlier than non-White participants (p = 0.01, Cohen’s d = -0.64). Bedtime was not different by ethnicity, first generation student status, or gender (p’s > 0.05).

### Salivary melatonin levels

We compared the impact of Dr. Teal’s Sleep Lotion with Melatonin and Essential Oils versus Dr. Teal’s Soothing Lavender Essential Oil Body Lotion as a placebo control. Participants provided saliva samples just before applying lotion (one hour prior to bedtime) and then 30 and 60 minutes later (at bedtime). Average melatonin levels on the placebo control night did not differ by race, ethnicity, first generation student status, gender, exercise, or caffeine (p’s > 0.05). Repeated measures 2 x 3 ANOVA showed main effects of treatment, F (1, 35) = 330.92, p < 0.001, ηp^2^ = 0.91; sampling time, F (2, 70) = 241.75, p < 0.001, ηp^2^ = 0.87; and the interaction effect, F (2, 70) = 191.48, p < .001, ηp^2^ = 0.85. On the placebo control lotion night, melatonin levels rose from a median of 34.1 pg/mL an hour before bedtime to 54.8 pg/mL at bedtime, as expected due to circadian effects (Figure 2). The application of Dr. Teal’s Sleep Lotion dramatically increased participants’ salivary melatonin levels up to ∼1000-fold compared to the placebo control (Figure 2).

**Figure 2.**
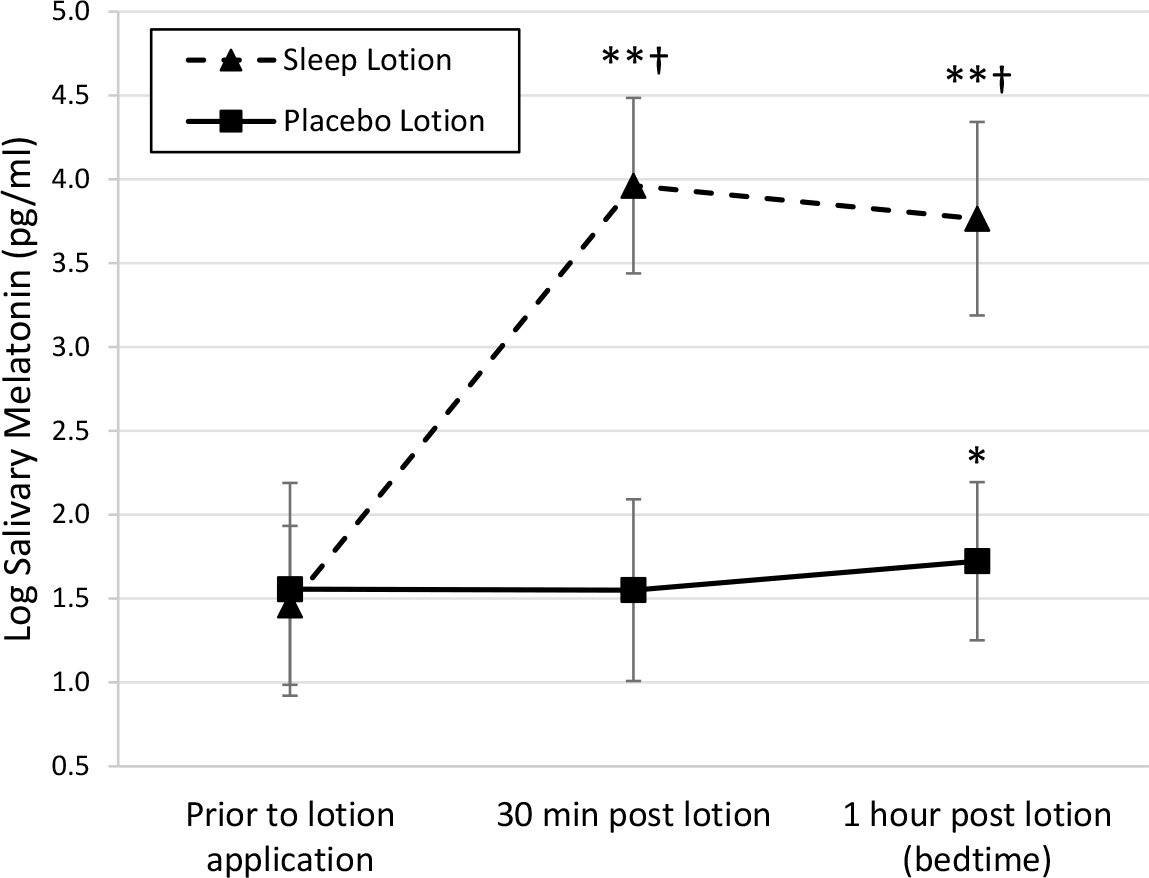
The impact of Dr. Teal’s Sleep Lotion with Melatonin and Essential Oils on salivary melatonin levels. Lotion was applied to hands and arms immediately after the first saliva sample, one hour prior to bedtime. Data were log transformed due to violations of normality; log_10_ of 10,000 pg/ml = 4. *p < 0.05 compared to prior to lotion application; **p < 0.001 compared to prior to lotion application; †p < 0.001 compared to placebo. n = 36 participants with all six timepoints.

### Impact of lotion on sleep quality

The morning after lotion application, participants completed a PSQI questionnaire modified to ask questions about the quality of the previous night’s sleep only. Instead of scores from 0 to 21, the possible scores on the modified questionnaire were 0 to 14. In an analysis of all participants, the average modified PSQI score on the night after placebo control lotion application was 3.25 ± 2.27, and on the night after sleep lotion application was 3.63 ± 2.54 (*p* > 0.05). We were particularly interested in the impact of lotion on sleep quality in the participants who were poorer sleepers, and thus we analyzed participants who scored at or above the median of 3.0 out of 14 on the modified PSQI after application of the placebo control lotion. In this subsample of 18 participants, the average modified PSQI score on the night after placebo control lotion application was 4.83 ± 1.72, and on the night after sleep lotion application was improved to 3.33 ± 2.22 (*p* = 0.02, Cohen’s d = 0.81).

### HPLC analysis of lotions

We measured the amount of melatonin in Dr. Teal’s Sleep Lotion with Melatonin and Essential Oils and Dr. Teal’s Soothing Lavender Essential Oil Body Lotion (placebo control) using HPLC. Melatonin standards at 10, 25, 50, 75, and 100 μg/mL displayed a distinct peak with a retention time of 12.2 seconds; the peak areas for these standards were used to create a linear calibration curve. Samples extracted from Dr. Teal’s Sleep Lotion had a matching peak and were determined to contain 46.5 ± 1.7 μg melatonin/mL extract. To calculate the percent recovery in our extraction method, we spiked an additional 50 μg melatonin/mL into the same amount of Dr. Teal’s Sleep Lotion. We recovered an additional 9.9 ± 1.1 μg melatonin/mL extract in our spiked samples compared to the non-spiked samples. Thus, our extraction method recovered 20% of the spiked melatonin, and presumably a similar percent from the original lotion sample. Taking this recovery rate into account, we estimate that Dr. Teal’s Sleep Lotion contains 2.4 ± 0.1 mg melatonin/g lotion for a 0.24 ± 0.01% formulation (g/100 g lotion). Given that our participants were supplied with 7 g of lotion and instructed to use it all, their dose was approximately 17 mg transdermal melatonin. Dr. Teal’s Soothing Lavender Essential Oil Body Lotion (placebo control) did not contain any detectable melatonin.

## Discussion

Average sleep duration, bedtime, and quality of sleep in our sample were similar to a previous study in a university population (Lund et al., 2010), although that study found that exercise frequency was not related to PSQI scores. However, our results that exercisers had higher sleep quality than non-exercisers would add to the weight of the evidence for this relationship (Wang & Biro, 2021). Our finding that White participants went to bed 42 minutes earlier than non-White participants is intriguing. Although reported typical total sleep time was not different by race in our study, the shift of sleep to a later clock time in non-White participants could compromise optimal function. Such delayed sleep phase has been associated with depression in some populations (Combs et al., 2021). Our data was collected by self-report; future work to confirm this finding would benefit from more objective actigraph data.

Previous studies have examined over-the-counter melatonin oral formulations (liquids, pills, tablets, and gummies) and found startling inconsistencies in the amount detected, up to almost five times the dosage listed on the label (Cohen, Avula, Wang, Katragunta, & Khan, 2023; Erland & Saxena, 2017). Dr. Teal’s lotions, bubble baths, and sprays have no information on the labels about the amount of melatonin in the products, how much of the products consumers should use, or how often consumers should use them. This includes Dr. Teal’s products specifically designated for children (https://www.drteals.com/products/?product-type=kids). Our data show extremely high salivary melatonin levels in response to sleep lotion application, and it complements appeals for increased regulation of this hormone in the United States (Grigg-Damberger & Ianakieva, 2017) and research on long-term safety (Li et al., 2022).

## Conclusion

We have shown here that Dr. Teal’s Sleep Lotion, a very common non-prescription transdermal formulation with no specific dosage listed on the label, contains 0.24 ± 0.01% melatonin that was readily absorbed up to ∼1000-fold higher than physiological levels. In a small undergraduate sample, this lotion also improved sleep in those with poor sleep quality. Consumers should be aware of the potency of such over-the-counter products.

## Data Availability

All data produced in the present study are available upon reasonable request to the authors.

## Acknowledgements

We thank the University of Redlands Faculty Research Grant program for supporting this project, and N. Olson for her contributions to the initial idea.

